# Electricity Free LAMP: A Protocol for Loop-Mediated Isothermal Amplification Using an Exothermic Chemical Reaction and Lyophilized Reagents

**DOI:** 10.64898/2026.11.06.26365637

**Authors:** Krishna Grandhe, Timothy E. Riedel

## Abstract

Loop-mediated isothermal amplification (LAMP) offers a promising alternative to PCR for low-resource settings, but practical barriers remain. For example, there is often the need for reliable electricity, precision heating equipment, and cold-chain storage of reagents. Here, we describe a rudimentary but functional platform that integrates (i) a non-electric, chemically driven water bath capable of sustaining LAMP-relevant temperatures and (ii) lyophilized LAMP assays compatible with simple sample inputs such as diluted saliva or oral rinse. We demonstrate successful LAMP amplification under multiple heating configurations, including a thermos-based chemical bath and field-mimicking setups. This work provides proof-of-concept for a low-cost, portable molecular diagnostic workflow that minimizes infrastructure requirements while maintaining analytical performance.

## Introduction

Loop-mediated isothermal amplification (LAMP) is a nucleic acid amplification technique that enables rapid, highly specific DNA or RNA detection under constant-temperature conditions, typically between 60–65 °C (Notomi et al., 2000). Unlike polymerase chain reaction (PCR), LAMP does not require thermocycling instrumentation, instead relying on strand-displacing DNA polymerase and a set of four to six primers recognizing distinct regions of the target sequence. This simplified thermal requirement, combined with high amplification efficiency and tolerance to crude sample matrices, has positioned LAMP as a leading candidate for decentralized and point-of-care molecular diagnostics (Moehling et al., 2021).

Despite its advantages, the widespread deployment of LAMP in low-resource settings continues to be limited by infrastructure constraints. Stable thermal incubation, even at a single temperature, typically requires powered heating devices. Additionally, many molecular reagents require freezing or refrigeration, necessitating cold-chain logistics. According to the World Health Organization (WHO), over one in four healthcare facilities in low– and middle-income countries lack reliable access to electricity (World Health Organization, n.d.), significantly impairing diagnostic capacity. Furthermore, infectious diseases such as tuberculosis, dengue, and HPV-associated conditions remain disproportionately prevalent in regions where laboratory infrastructure is limited. These systemic barriers highlight the urgent need for electricity-independent, transportable molecular diagnostic systems.

To maximize accessibility in low-resource or field environments, the assay workflow was designed to minimize reliance on specialized instrumentation or laboratory infrastructure. The procedures can be performed using simple transfer methods such as capillary tubes or other fixed-volume transfer devices, reducing the need for precision liquid-handling equipment. Following collection and minimal preparation, the reaction mixture may be directly transferred into sealed amplification tubes preloaded with lyophilized or ready-to-use LAMP reagents. In terms of a heat source, non-electric heating strategies for isothermal amplification include passive thermal retention, combustion-based heating, phase-change material (PCM) systems, and chemically driven exothermic reactions. Passive and combustion-based methods are often limited by poor temperature control, environmental dependence, or safety concerns. PCM-based systems can stabilize temperatures effectively but frequently require engineered materials or specialized fabrication.

Chemically driven reactions provide a practical alternative because they generate heat internally without external power. Previously explored chemistries include calcium oxide hydration and magnesium-based alloy reactions, though many require PCM regulation to prevent overheating. In this work, heat was generated using the aluminum–sodium hydroxide reaction described later. This reaction was selected because it uses inexpensive, widely available materials and allows thermal output to be tuned through reagent concentration and reactant mass. Combined with the thermal buffering capacity of a water bath, this approach enables sustained incubation temperatures suitable for LAMP without requiring engineered phase-change materials or electronic control systems.

By addressing both thermal control and reagent stabilization, infrastructure-light amplification systems have the potential to expand access to molecular diagnostics in remote, field, and emergency environments. Reducing dependence on electrical power and refrigeration represents a critical step toward democratizing nucleic acid testing globally.

## Materials and Methods

### Non-Electric Heating System

The NEWB system utilizes a vacuum-insulated stainless-steel flask as the primary incubation chamber, allowing heat generated from the internal chemical reaction to be retained efficiently throughout the amplification period (Figure 1). Reaction tubes containing prepared LAMP mixtures were submerged in the heated water, with a string attached to the tubes for easy removal.

**Figure 1.**
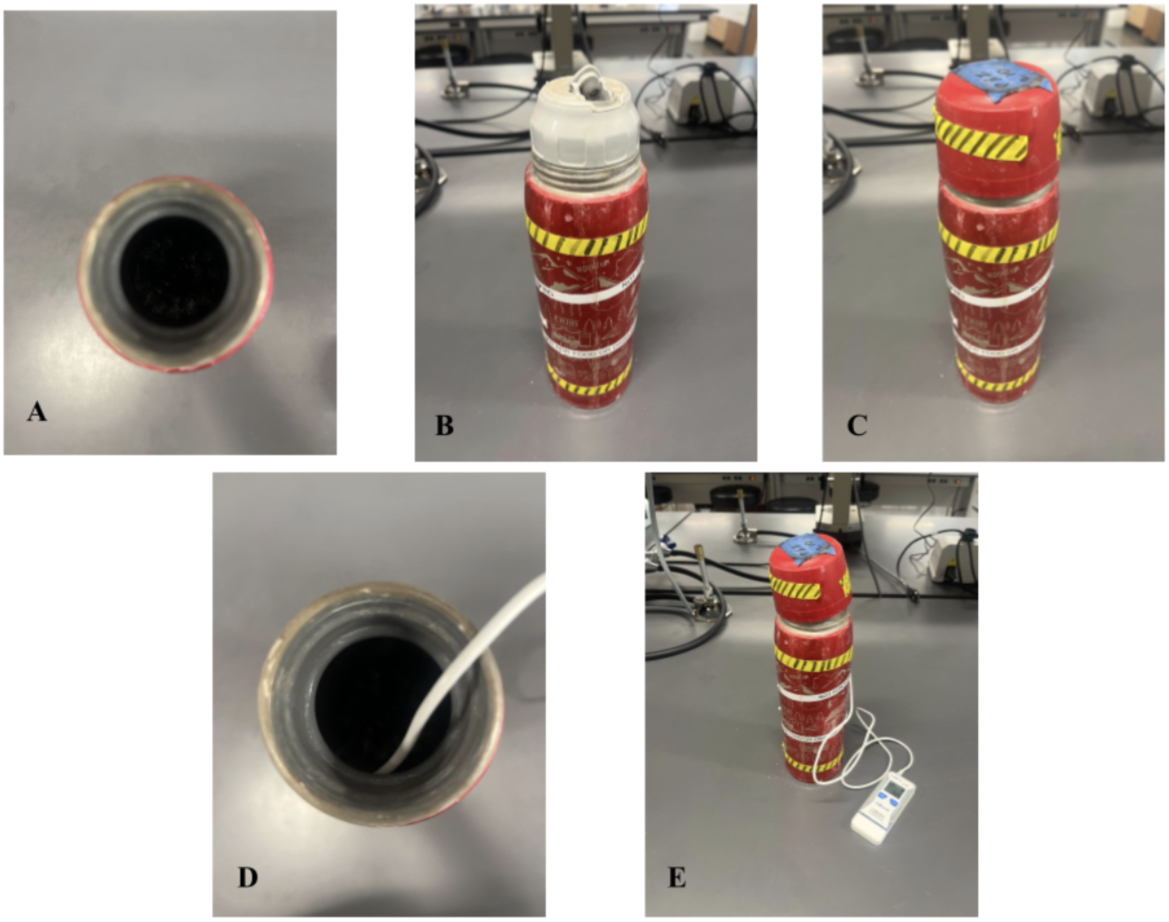
Assembly and operation of the non-electric water bath (NEWB) platform. The heater consists of an insulated vacuum flask containing the chemical heating reaction (A), a sealing lid that provides a controlled ventilation pathway for hydrogen gas generated during the reaction (B), an outer cap attached to reduce thermal loss and maintain incubation temperature (C), and a thermometer for real-time temperature monitoring (E). Note that the heater must remain vented during operation to release hydrogen gas produced from the chemical reaction.

To initiate operation, chemical reactants were added to the insulated chamber and allowed heat generation to begin prior to insertion of prepared amplification tubes. The inner lid was then secured while maintaining an open vent pathway to allow safe release of hydrogen gas generated as a byproduct of the reaction. Finally, the outer cap was attached to reduce heat loss and improve thermal retention during incubation. Temperature was continuously monitored using an external probe thermometer to characterize thermal performance throughout each experimental run. To thermally incubate LAMP reactions, tap water was heated using an exothermic chemical reaction between aluminum metal and sodium hydroxide in aqueous solution, with sodium chloride added to modulate reaction kinetics (Equation 1, Hurtubise et al., 2018).

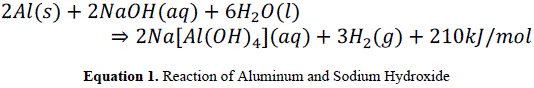

In this reaction, sodium hydroxide disrupts and dissolves the protective aluminum oxide layer naturally present on the metal surface, allowing the underlying aluminum to react with water. The oxidation of aluminum releases substantial thermal energy while producing sodium aluminate and hydrogen gas as byproducts. Sodium chloride was additionally included as an additive to promote disruption of the oxide layer and increase solution conductivity, facilitating more consistent reaction initiation and heat generation.

The system was intentionally designed for an initial water temperature of approximately 22 °C, such that the heat released would raise the solution into the LAMP-compatible temperature range, which was defined to be 63-67 °C. This was consistent with the ∼60–72 °C optimal performance range reported for Bst 2.0 WarmStart DNA Polymerase (Bst 2.0 WarmStart DNA Polymerase, NEB) and 57-67 °C range described for Bst Polymerase (Francois et al., 2011). The reaction mixture consisted of 4.06 g aluminum metal, 53 mL of 3 M NaOH, and 147 mL of water (total volume 200 mL), with 1.0 g NaCl (H-E-B Salt, H-E-B, LP, Austin, TX, USA) added to facilitate oxide layer disruption and promote sustained aluminum dissolution.

Aluminum used was in the form of aluminum foil (Boardwalk 7110BW Extra Standard, Miami, FL, USA) in rectangles of approximately 7 cm x 4 cm. These pieces were then crumpled into balls of diameter of 1cm (Figure 2). However, the amount was slightly varied so that exactly 4.06 g could be reached. 3M sodium hydroxide solution was prepared by combining tap water with sodium hydroxide pellets (Fisher Scientific S320-500, Fair Lawn, NJ, USA), and mixing with a lab stir plate until completely dissolved. The reaction was completed in a 1.03 L vacuum insulated stainless steel bottle (Zojirushi SJ-JS10RA, Osaka, Japan). The reactants were swirled by hand in the bottle for approximately 10 seconds to promote reaction. Reaction tubes were submerged in the water bath and incubated without active temperature control. Temperature profiles were collected using a digital logging thermometer (Elitech Data Logger LogEt 5TE, San Jose, CA, USA).

**Figure 2.**
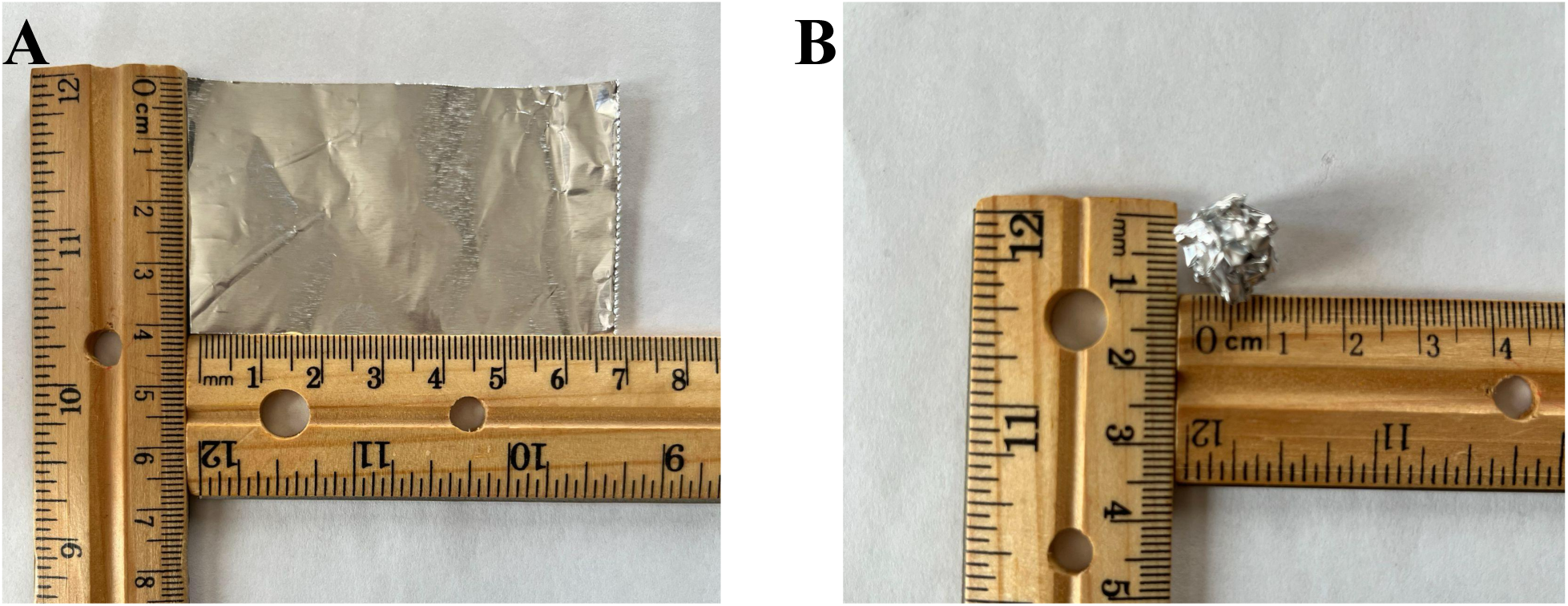
Amount of aluminum foil used to drive exothermic chemical reactions. A) The initial size of the foil square used to create a crumpled ball of foil. B) Approximate size of crumpled ball version added to chemical reactions in vacuum flask.

### LAMP Reagents and Primer Preparation

NEWB was tested against three previously validated LAMP reactions (Hill, Saitiew, Parida) (Table 1). The primers were prepared as a 10× primer solution. For lyophilization, approximately 16.5 µL of the 10× primer mix was aliquoted per tube and lyophilized. Prior to use, primers were reconstituted with 165 µL of molecular grade H_2_O (molH_2_O), yielding a working concentration suitable for 25 µL LAMP reactions. In some trials, DI water was replaced with an oral rinse solution to simulate field-compatible inputs.

**Table 1:**
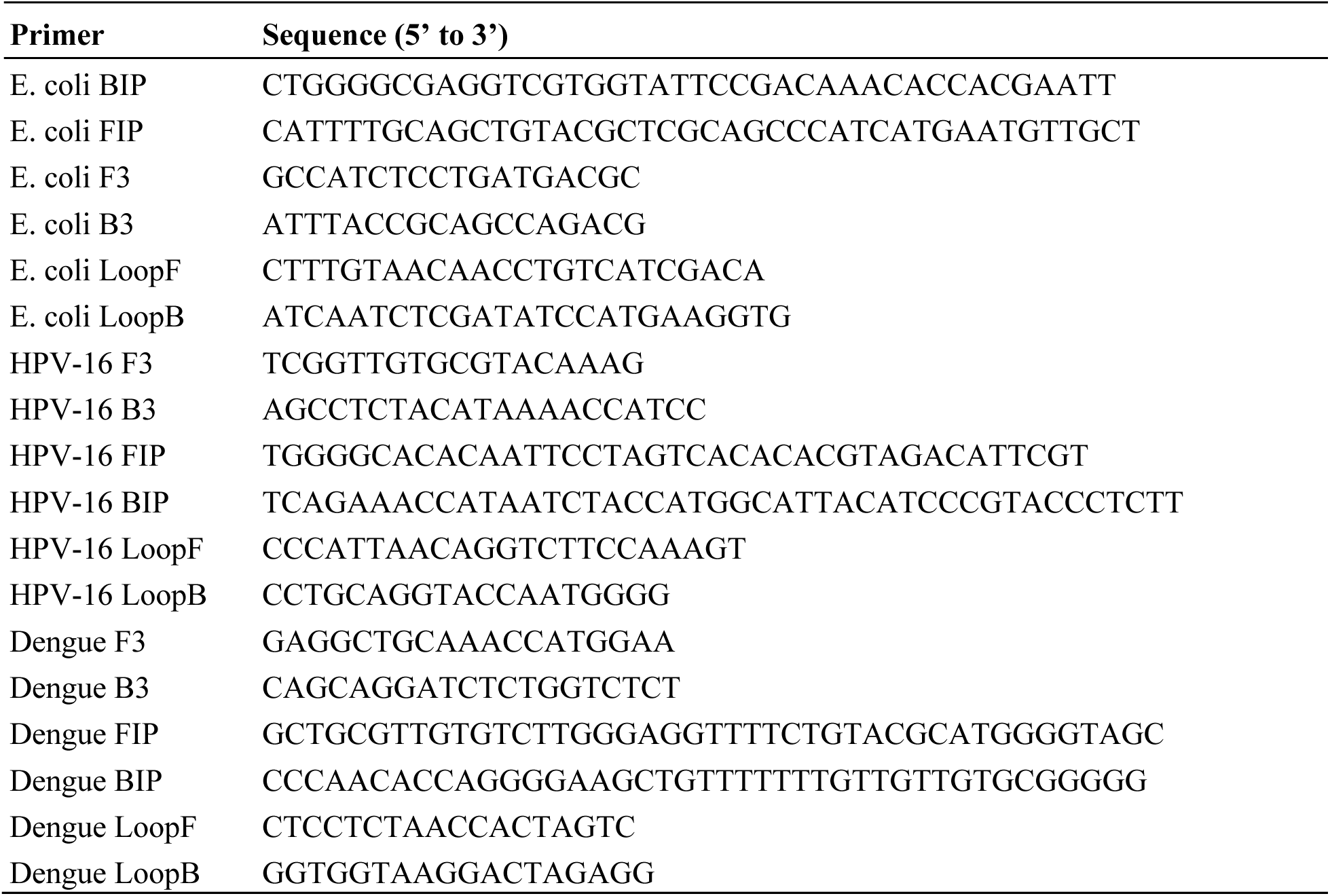
*E. coli* (Hill et al., 2008), HPV-16 (Saetiew et al., 2011) and Dengue Virus (Parida et al., 2005) LAMP primers used in this study.

Each experimental run consisted of four to six 25 µL reactions, with two to three no-template controls (NTCs; 1 µL DI water or oral rinse) and an equal number of positive controls (1.5 µL template, ∼10^4^ copies/μL). Note that although most experiments were carried out with a PTC of 10^4^ copies/μL, 10^3^ was also run with similar results.

### Sample Input

To model non-invasive human sampling, oral rinse was simulated using a 15:1 ratio of bottled water (Aquafina) to saliva surrogate (D’Souza et al., 2005; Z. Li et al., 2022). This dilution was chosen to balance inhibitor tolerance with target detectability.

### Image Analysis

Line profiles were generated in ImageJ (Schneider et al., 2012), and peak intensities were extracted from gray value measurements for downstream analysis.

### LAMP Incubation and Readout

Reactions were incubated for up to 50 minutes. At the current stage of validation, fluorescent LAMP (fluoro-LAMP) was used as the primary readout modality. Fluorescence was visualized using a P51 fluorescence viewing box (MiniPCR Bio, Cambridge, MA), which enabled reliable discrimination between amplified and non-amplified reactions. Checks were done with the unaided eye through the viewing box every 10 minutes. For the purposes of this paper, successful detection is defined as clear temporal separation between positive controls and negative controls. A positive reaction was indicated by a higher intensity of brightness relative to negative controls. For colorimetric LAMP, reactions were incubated for an equal amount of time. The positive reaction was defined to be a yellow tube, which negative reaction was defined to be a pink tube.

### Statistical Analysis and Tools

Statistical analyses were selected according to the structure of each dataset. Temperature-performance data were summarized using descriptive statistics, including peak temperature, time within the LAMP-compatible temperature range, and cooling behavior over the incubation period. Colorimetric LAMP outcomes were treated as ordinal categorical data, with pink indicating no amplification, orange indicating partial/intermediate amplification, and yellow indicating full amplification. Agreement between NEWB and conventional water bath conditions was summarized using percent agreement and weighted Cohen’s kappa. For endpoint diagnostic comparisons, color outcomes were additionally binarized with yellow scored as positive and pink/orange scored as non-positive, and paired binary outcomes were compared using McNemar’s exact test. Time-to-positivity (TTP) was defined as the first time point at which a visible yellow color change occurred and was summarized descriptively at the replicate level.

Where continuous fluorescence or threshold-cycle values were compared between positive and negative reactions, two-sample t-tests were used unless otherwise noted.

## Results

### Thermal Performance of the Non-Electric Water Bath (NEWB)

The thermal performance and stability of the NEWB heater were characterized under laboratory bench-top conditions. During each trial, the water temperature inside the heater was monitored for at least two hours using a submerged digital thermometer while the system was operated at room-temperature ambient conditions. Across three trials, the water bath reached 63 °C in approximately 23 minutes and maintained the LAMP-relevant range of 63–67 °C for an average of 80.3 minutes (Figure 3).

**Figure 3.**
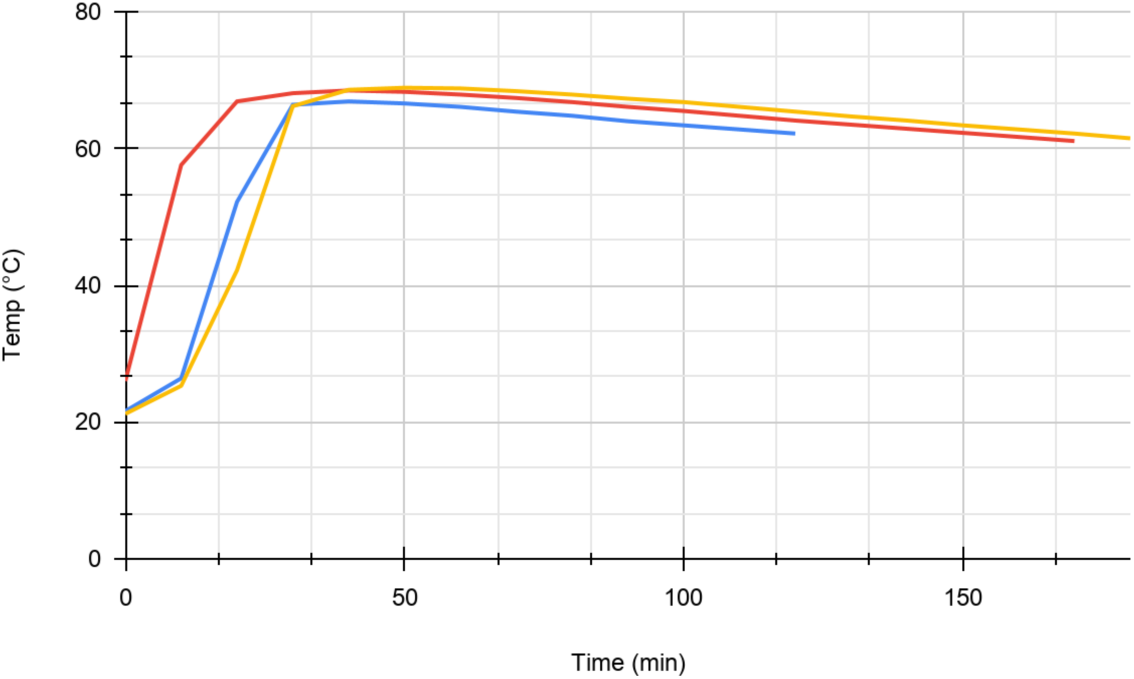
Temperature stability of NEWB over time. Three separate experimental runs were monitored with a logging thermometer for over two hours. Once achieving peak temperature, heat loss from the water is approximately 4 degrees Celsius for over 80 minutes.

The NEWB system was then tested outside the lab in a field where the ambient temperature was measured at 15 °C and the heater achieved a stable temperature of 61.5 °C (Figure 4).

**Figure 4.**
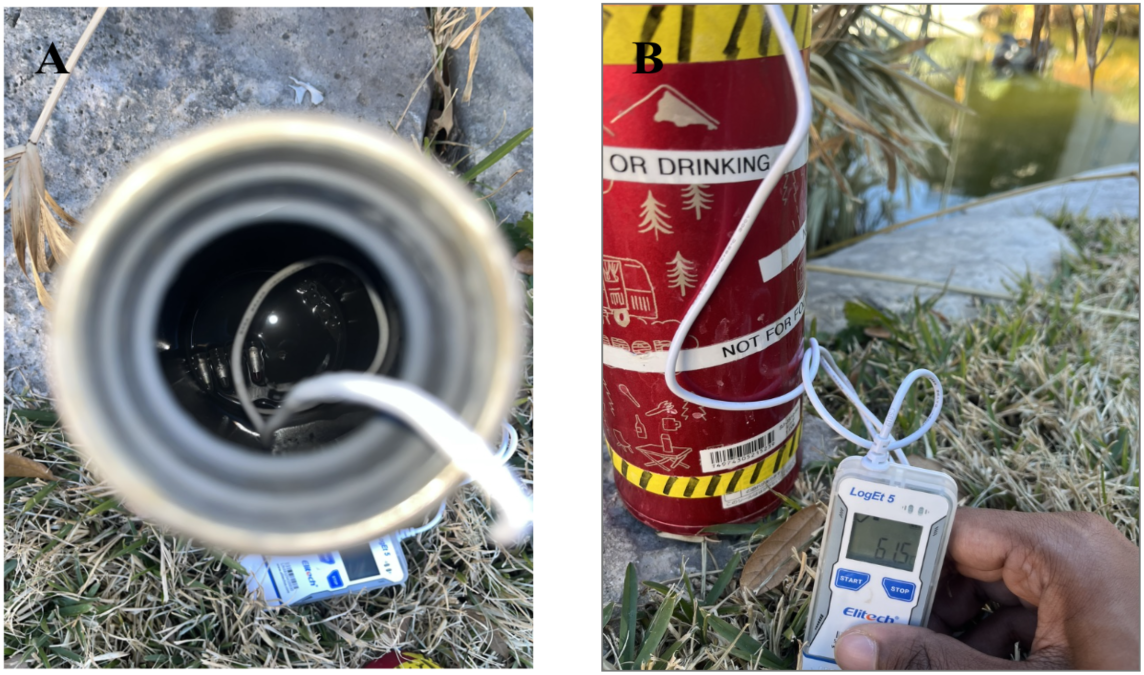
Example setup of NEWB in an outside environment. Panel A shows the assay in NEWB. Panel B shows the setup in an outside environment. The ambient temperature was recorded to be 15 °C.

### LAMP Amplification in Water Bottle Setup

To validate that the NEWB setup supports LAMP amplification, reactions were performed within the NEWB prewarmed water and compared to side-by-side reactions in a lab-grade electric water bath. Two LAMP primer sets, one designed to detect Dengue Virus (DEN-1) and one designed to detect HPV (strains 16 and 18) were tested together. Positive control reactions produced detectable signals while no-template controls either remained negative or amplified at least 20 minutes after the positives (Table 2).

**Tables 2a and 2b.**
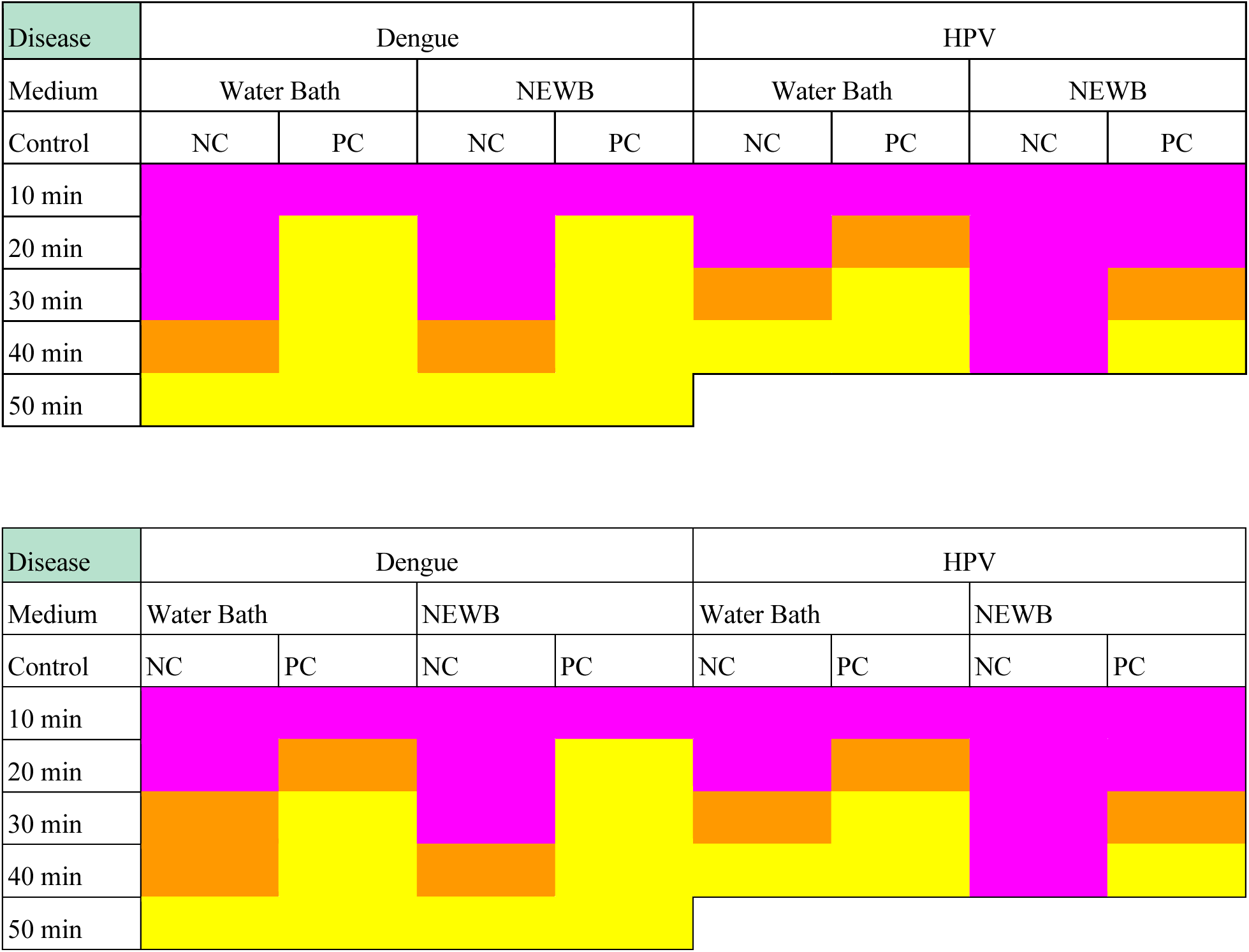
Comparison of LAMP assay performance in a lab-grade water bath versus NEWB. Pink represents no amplification, orange represents partial amplification, and yellow represents full amplification. Primer sets for Dengue and HPV-16 were tested. Out of the 36 color scores, 34 were identical over the replicates.

| Disease | Dengue |  |  |  | HPV |  |  |  |
| Medium | Water Bath |  | NEWB |  | Water Bath |  | NEWB |  |
| Control | NC | PC | NC | PC | NC | PC | NC | PC |
|---|---|---|---|---|---|---|---|---|
| 10 min |  |  |  |  |  |  |  |  |
| 20 min |  |  |  |  |  |  |  |  |
| 30 min |  |  |  |  |  |  |  |  |
| 40 min |  |  |  |  |  |  |  |  |
| 50 min |  |  |  |  |  |  |  |  |
| Disease | Dengue |  |  |  | HPV |  |  |  |
| Medium | Water Bath |  | NEWB |  | Water Bath |  | NEWB |  |
| Control | NC | PC | NC | PC | NC | PC | NC | PC |
| 10 min |  |  |  |  |  |  |  |  |
| 20 min |  |  |  |  |  |  |  |  |
| 30 min |  |  |  |  |  |  |  |  |
| 40 min |  |  |  |  |  |  |  |  |
| 50 min |  |  |  |  |  |  |  |  |

For direct comparison of heating modalities, color outcomes were assigned ordinal scores of pink = 0, orange = 0.5, and yellow = 1. Agreement between water bath and NEWB was summarized using percent agreement and weighted Cohen’s kappa. Endpoint diagnostic agreement was additionally assessed after binarizing outcomes as yellow = positive and pink/orange = non-positive. For dengue, NEWB showed strong percent agreement with the conventional water bath, with 18 of 20 paired observations matching exactly and strong ordinal agreement by weighted

Cohen’s kappa (κ = 0.90). Binary endpoint comparison showed no significant difference between heating conditions (McNemar’s exact test, p = 1.00). Time-to-positivity (TTP), defined as the first time point at which a visible yellow color change occurred, was similar between methods, with only one replicate showing a 10-minute difference.

For HPV, agreement between heating modalities was lower, with 8 of 16 paired observations matching exactly and fair-to-moderate ordinal agreement (weighted κ = 0.35). Water bath reactions progressed to yellow earlier than NEWB in several paired observations; however, binary endpoint comparison did not reach statistical significance (McNemar’s exact test, p = 0.125). Despite this modest delay, NEWB still generated a clear diagnostic window of at least 20 minutes between positive and negative reactions, supporting its practical utility for endpoint LAMP detection.

### Lyophilized LAMP Primers in NEWB

To test whether lyophilized LAMP can be performed outside the lab, both lyophilized and non-lyophilized E. coli primer sets were tested in the NEWB (Figure 5).

**Figure 5.**
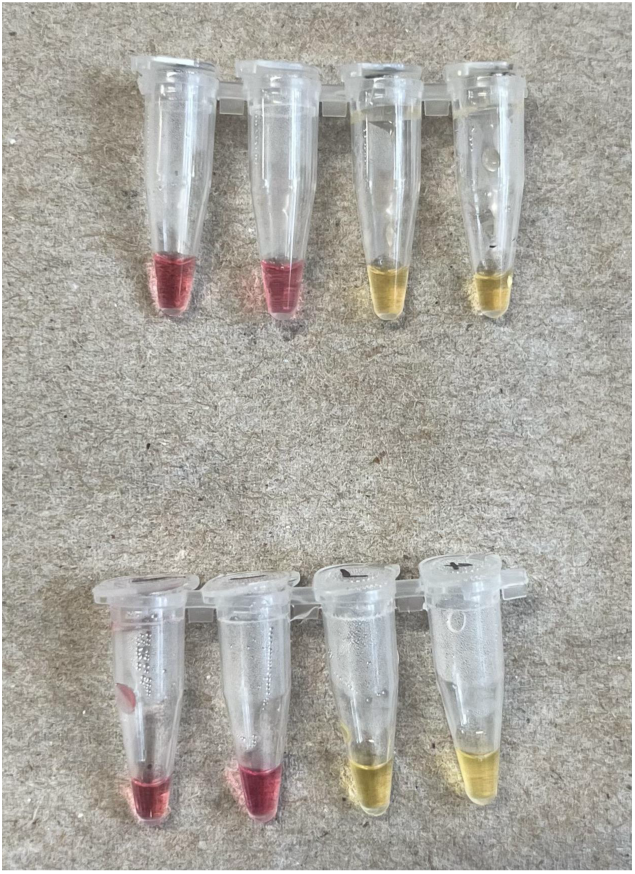
Comparison of LAMP in a stand. The top set of tubes used standard primers, while the bottom set used lyophilized primers. The left two tubes of each set were negative controls, with the remaining being positive controls. After 40 minutes, results were recorded.

### Lyophilized Master Mix and Lyophilized Primers

To expand the low-resource usefulness of NEWB, this study explored the use of lyophilized enzymes in addition to primers incubated in the NEWB setup. Colorimetric master mixes are not typically amenable to lyophilization (personal communication). LyoPrime (NEB, Ipswich, Massachusetts) was utilized with the in-house lyophilized primer sets and incubated in NEWB. The fluorescent end-point performance validates this system can discriminate between negative and positive controls (Figure 6).

**Figure 6.**
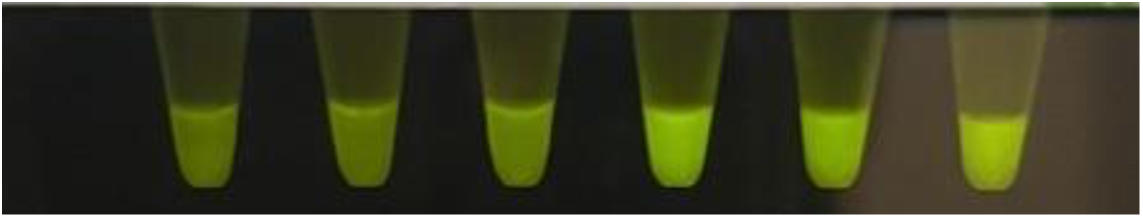
LyoPrime *E. coli* Assay viewed through P51 fluorescent visualizer system. Left three tubes are negative controls, with the right three being positive controls. A positive reaction is indicated by a higher relative brightness. The unedited picture was taken at the 30-minute mark.

To ensure there is a quantitative difference between fluorescent intensity between the positive and negatives, we analyzed the image with computer software (ImageJ) that converts brightness in grey values (Figure 7).

**Figure 7.**
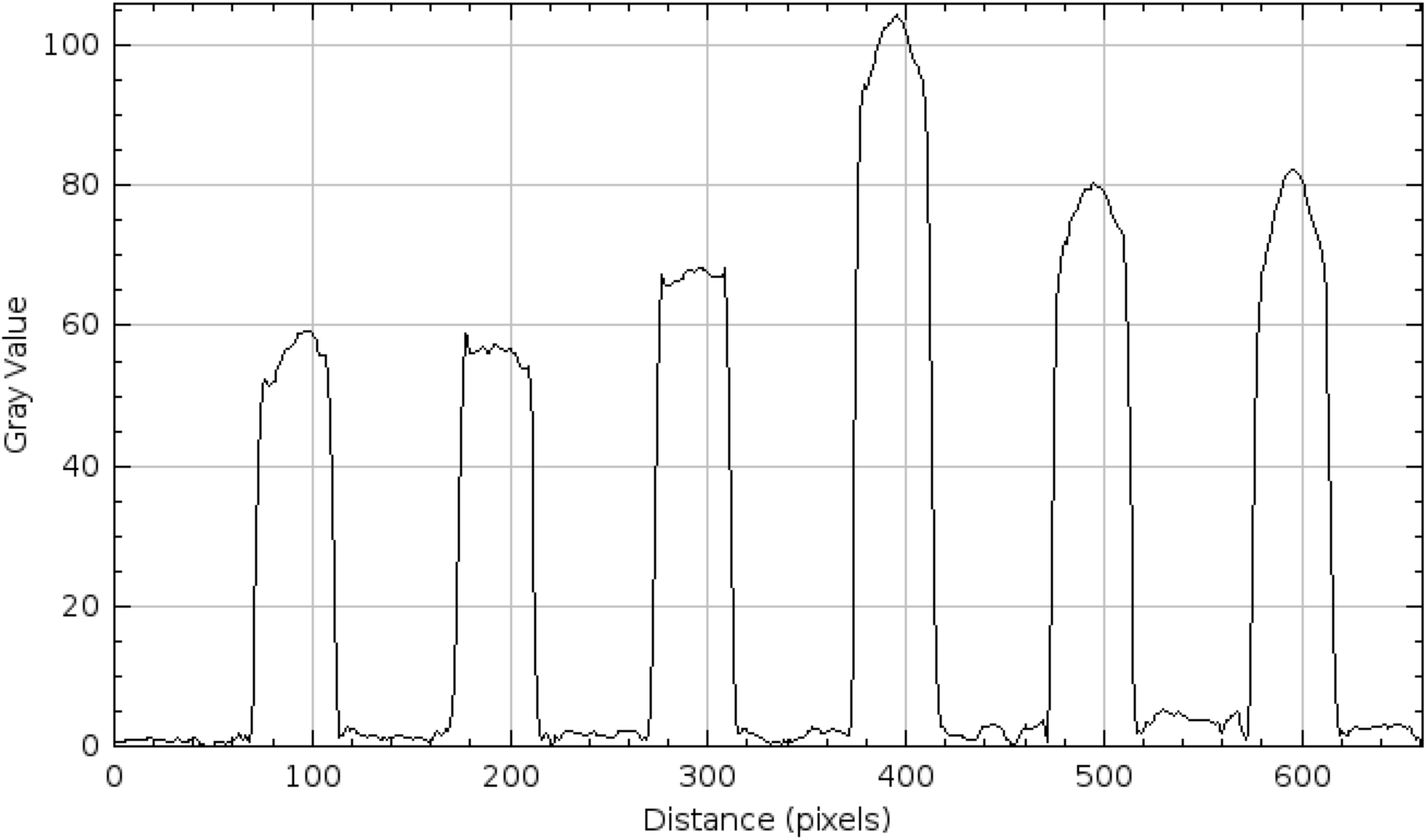
Quantitative analysis of image intensity following LAMP amplification. The endpoint image (Figure 6) was processed in ImageJ using rolling-ball background subtraction (50 px radius), and a line profile estimated to be the middle of the tubes was extracted to obtain pixel intensity as a function of position.

Peak intensities were averaged per tube (n = 3 per group). Positive samples (mean = 77.72, SD = 11.96) were higher than negative controls (mean = 56.82, SD = 6.30). Comparisons using a Welch’s t-test show (t = −2.68, p = 0.074).

The lyophilized *E. coli* assay was further characterized using a qPCR machine set to hold a constant temperature of 65 and take a fluorescent reading every 70 seconds. Positive controls (Figure 8 blue, red) demonstrated rapid amplification with a mean threshold (150 RFU) time of 15.5 ± 0.7 minutes, whereas negative controls (Figure 8 orange, green) amplified substantially later (mean ≈ 48 ± 10 minutes). The separation between groups was highly significant (two-sample t-test, p < 0.01), with a difference of approximately 32.5 minutes.

**Figure 8.**
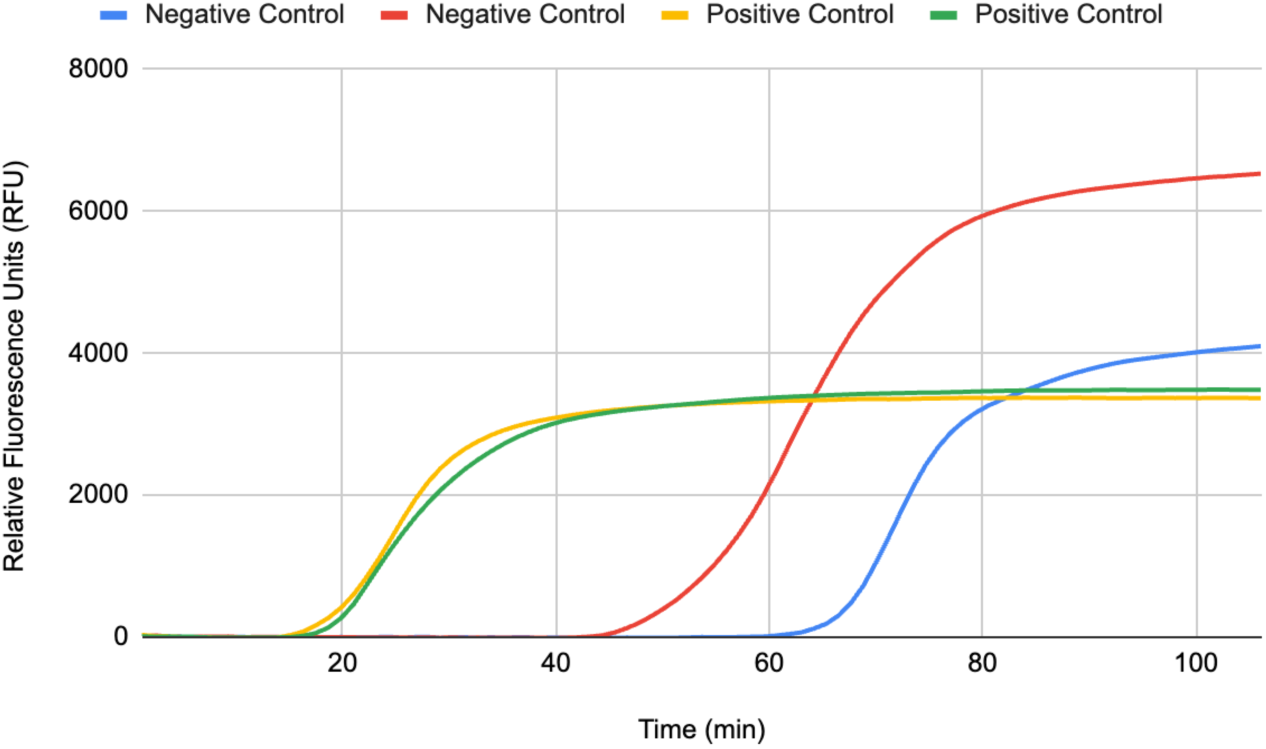
Fluorescence versus time of the lyophilized *E. coli* assay incubated and monitored in a qPCR machine.

### Oral Rinse

When replacing DI water with oral rinse as outlined in the protocol, fluorescent amplification via qPCR machine was very similar to using the standard DI water (Figure 9). Positive controls amplified at 20 minutes, with negative controls amplifying at 95 minutes. This results in a 75-minute diagnostic period.

**Figure 9.**
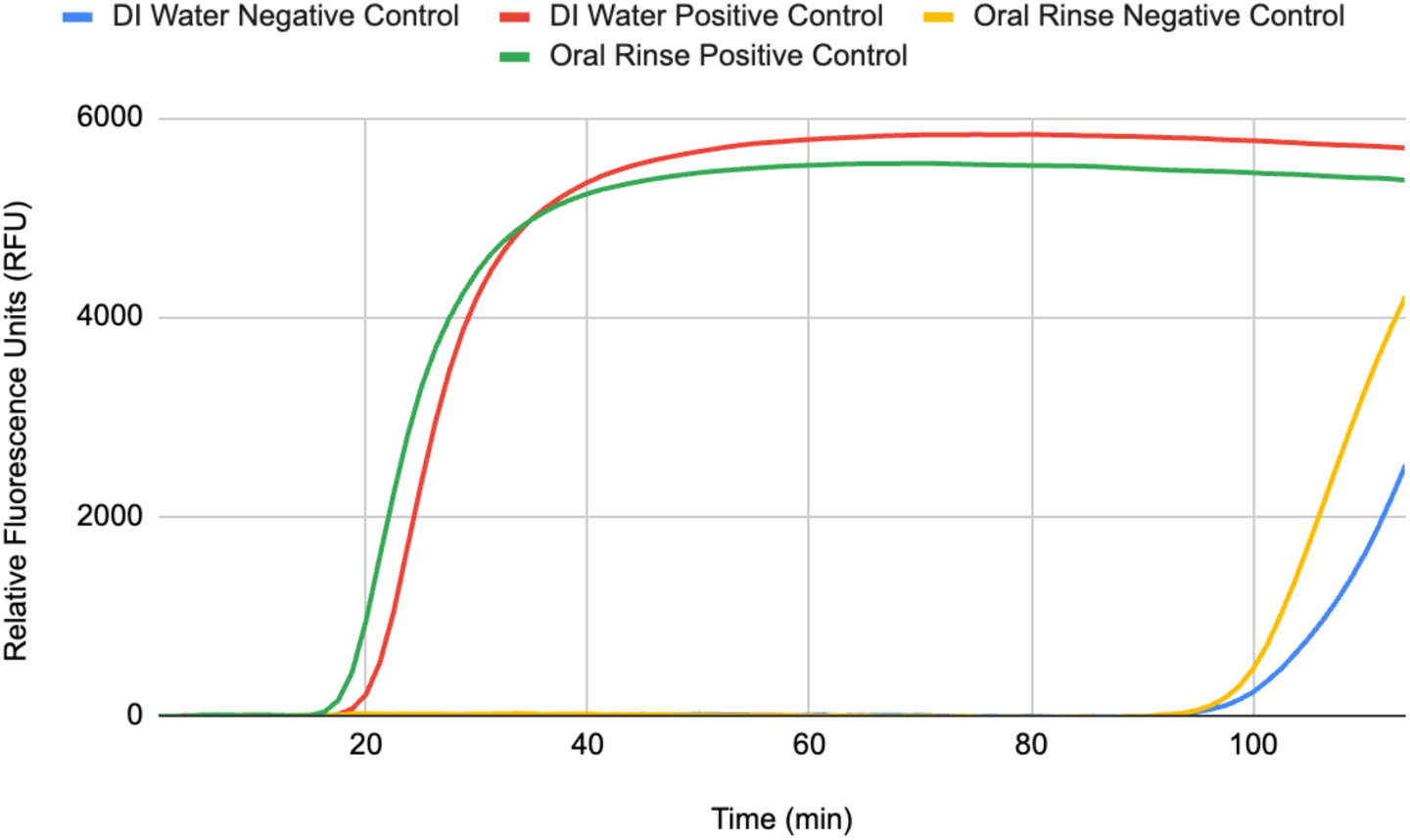
Quantitative comparison of DI water versus oral rinse in protocol.

Specifically, paired comparison of fluorescence values across all 91 cycles demonstrated no statistically significant difference between DI water negative control and Oral Rinse negative control, nor between DI water positive control and Oral Rinse positive control (paired t-test, p > 0.05 for both comparisons). The cycle-by-cycle differences were small relative to overall signal amplitude, and amplification onset and plateau phases occurred at comparable cycles within each replicate pair. These results confirm strong intra-condition reproducibility and demonstrate that both sets of pairs exhibit equivalent amplification behavior.

Overall, lyophilized *E. coli* primers successfully supported LAMP amplification following rehydration. Positive control reactions consistently amplified earlier than negative controls, demonstrating both primer integrity and reaction specificity after lyophilization and storage.

## Discussion

This study demonstrates that LAMP-based diagnostics can be carried out in a non-electrical setting. An example SOP based on this study is provided (Appendix A). Lyophilized LAMP primers were combined with a non-electric, chemically generated heat source to perform nucleic acid amplification without conventional laboratory infrastructure. For example, the NEWB system was used successfully on the bank of a creek (Figure 4).

Prior studies have investigated portable incubation strategies for LAMP and other isothermal amplification reactions, including disposable chemical warmers (Hatano et al., 2010) and compact chemical heaters (Shah et al., 2015). Many electricity-free approaches have relied on exothermic chemical reactions coupled with engineered phase-change materials (PCMs) to buffer heat release and maintain the reaction near the desired amplification temperature (R. J. Li et al., 2021) (Table 3). While such approaches represent important advances toward portable molecular diagnostics, they may require specialized fabrication or device components. In contrast, chemically driven heating systems can provide tunable thermodynamic behavior, allowing incubation temperatures to be adjusted through reagent quantities or reaction conditions. This flexibility is particularly advantageous because LAMP temperature requirements vary with assay as demonstrated by E. coli detection at 66 °C (Hill et al., 2008), high-temperature LAMP formats at 68–72 °C (Simões et al., 2026), and a SARS-CoV-2 LAMP assay at 58 °C (Luo et al., 2022).

**Table 3.** Comparison of Non-electric Isothermal Amplification Setups.

| **Feature** | **PCM Thermos Incubator (Singleton et al., 2014)** | **Energetic Phase-Change Heater (EPCM) (Li et al., 2021)** | **NEWB (This Study)** |
|---|---|---|---|
| **Heating mechanism** | Exothermic chemical heater coupled to insulated thermos | Mg–Fe alloy + water exothermic reaction | Aluminum + NaOH + water exothermic reaction |
| **Temperature control strategy** | Palmitic acid phase-change material stabilizes incubation temperature | Energetic phase-change material (PCM combined with exothermic fuel) buffers heat output | Thermal buffering of water bath combined with controlled reaction stoichiometry |
| **Nominal incubation temp.** | ~61.5 °C ± 1.5 °C | ~60–65 °C | Tunable (~63–67 °C demonstrated) |
| **Ramp-up time** | Rapid (<2 min reported) | Rapid (~3 min reported) | ~20–25 min |
| **Thermal plateau duration** | ~60 min | ≥30–60 min | ~80 min within target temperature range |
| **Phase-change material used** | Yes (palmitic acid) | Yes (PCM composite) | No |
| **Material complexity** | Moderate (PCM + metal holder + thermos) | Moderate–high (engineered EPCM composite) | Low (commodity chemicals and insulated container) |
| **Manufacturability / scalability** | Moderate | Moderate | High due to simple materials and assembly |
| **Detection modality demonstrated** | Colorimetric / lateral flow HIV detection | RT-LAMP colorimetric SARS-CoV-2 detection | Fluorescent and colorimetric LAMP validation |
| **Reported sensitivity (LOD)** | Comparable to laboratory LAMP assays | Detects viral RNA at clinically relevant levels | ~10<sup>3</sup> copies per reaction (demonstrated) |
| **Primary limitations** | Requires PCM preparation and dedicated device components | Requires engineered EPCM heater materials | Slower ramp-up and gas-generating reaction requiring ventilation |

The heating strategy described in this study examines the feasibility of direct chemical heating without PCMs. Some potential advantages are, First, it relies on commercial off-the-shelf chemical reactants and common insulated containers, eliminating dependence on specialized phase-change materials or engineered thermal substrates. Second, the system’s thermal output can be modulated by adjusting reagent quantities, enabling accommodation of diverse isothermal amplification temperature requirements. Experiments were run at an intended starting water temperature of 22 °C. However, the initial quantities of Al and NaOH can be manipulated such that it reaches around 67 °C consistently (Equation 2).

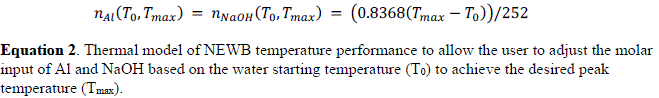

where *T_0_* is the initial water temperature. The coefficients were derived from observed *ΔH*^∘^ values from the NEWB in this study (Figure 3), which was approximately-252 kJ/mol on average. Third, the absence of electronic components reduces manufacturing complexity, improves transportability, and eliminates reliance on external power sources or batteries. Collectively, these features enhance scalability and accessibility, particularly for deployment in low-resource or emergency settings.

A crude cost analysis suggests that the chemical heating system can be assembled for under $4 per reaction (Table 4). Note, it was assumed 12 reactions can be tested per run.

**Table 4.** Estimated cost of running LAMP reactions in the NEWB setup. Note that the initial cost of the Thermos ($30) was not factored into the costs in the table and we assumed 12 reactions can be tested per run.

| Cost Category | Per Reaction Cost |
|---|---|
| Chemical heater (NaOH + Al + NaCl) | \$0.19 |
| LAMP reagent master mix | \$2.52 |
| Primers | \$1.23 |
| Molecular H2O | \$0.02 |
| Total (all components) | \$3.96 |
| Fixed Cost: Thermos | \$30.00 |

A preliminary cost comparison indicates that the material expenses associated with the NEWB heating platform are comparable to previously reported electricity-free incubation systems. The chemical heating components of the NEWB system (NaOH, aluminum, and NaCl) were estimated to cost approximately $0.19 per reaction, assuming 12 reactions are performed simultaneously. In comparison, the heater reported by Singleton et al. estimated chemical heating material costs of approximately $0.06 per test. Although the NEWB heating components are modestly more expensive on a per-reaction basis, both systems contribute less than one dollar per assay, indicating that the heating platform contributes minimally to the overall cost of electricity-free molecular diagnostics.

Several practical insights emerged. First, the chemical water bath produced sufficiently stable temperatures despite lacking feedback control in both interior (Figure 3) and exterior environments (Figure 4). Due to the inherent robustness of LAMP chemistry, reactions for two different primer sets performed similarly when directly compared to reactions in a lab grade water bath (Table 2). Despite a time delay for the reactions incubated in NEWB, they all still generated a clear difference between true positives and true negatives, supporting its practical utility for endpoint LAMP detection. Second, lyophilization of primers did not impair amplification, supporting the feasibility of room-temperature reagent storage (Figure 5). Third, although fluorescent detection was used for validation (Figures 6 and 7), transitioning to colorimetric lyophilized LAMP would further simplify the workflow by eliminating the need for complicated optical components.

Limitations of this work include incomplete integration of the colorimetric lyophilized master mix at the time of writing, as well as the need for broader validation across users, assay formats, and environmental conditions. Because NEWB relies on passive chemical heating rather than active electronic feedback, its temperature profile may be influenced by ambient temperature, flask geometry, insulation performance, reagent mixing, and starting water temperature. As a result, the system will be less precise than a laboratory-grade heat block or water bath, may heat more slowly, and will require calibration for each flask or field configuration before use (Equation 2).

Additional practical considerations include chemical waste handling and hydrogen gas generation. The aluminum–sodium hydroxide reaction produces an alkaline waste stream that must be disposed of appropriately and releases hydrogen gas as a byproduct; for example, 4.06 g of aluminum is expected to generate approximately 5.1-5.5 L of hydrogen gas under atmospheric conditions. These factors emphasize the need for venting, avoidance of ignition sources, and operation in a well-ventilated setting.

Future work should focus on development of colorimetric lyophilized LAMP, which would eliminate the need for fluorescence viewing equipment altogether and further simplify result interpretation. Additional efforts should include developing consistent reagent packaging, validating long-term reagent stability, and expanding testing to additional LAMP-diagnosable targets.

## Conclusions

In principle, nucleic acid targets compatible with LAMP amplification could be adapted to this platform after assay-specific validation. This enables molecular diagnostics to be performed in a portable, low-cost, and electricity-free manner. Because the system relies on chemically generated heat, lyophilized LAMP reagents and NEWB together could be stored, shipped, mailed, or sold without refrigeration or operational power requirements. This opens opportunities for at-home testing, remote clinical screening, environmental monitoring, and rapid deployment during outbreaks or emergencies.

## Supplementary Materials

The following supporting information is supplied as a PDF.

Example SOP

## Supporting information

Supplemental Materials, Example SOP

## Data Availability

All data produced in the present work are contained in the manuscript.

## Acknowledgments

This work was made possible thanks to donations from Bob and Cathy O’Rear and New England Biolabs. We would also like to thank the entire UT Austin CNS FRI DIY Diagnostic Research Lab (https://diystream.cns.utexas.edu/) community for their overall support and their assistance with sourcing the materials needed for this study, and Derek Bashe for his assistance with lyophilization during this project.

## Declaration of Generative AI and AI-assisted technologies in the writing process

The creation of this paper used ChatGPT to assist with language refinement and organization, identify relevant sources, and support the selection and calculation of various statistical analyses. All AI-generated content was reviewed and verified to ensure accuracy. ChatGPT was not used to generate experimental data or draw conclusions independently. The authors take full responsibility for the accuracy and integrity of the published work.

## Abbreviations

DNA: deoxyribonucleic acid
F3/B3: forward/backward outer primer
FIP/BIP: forward/backward inner primer
LAMP: loop-mediated isothermal amplification
LoopF/B: forward/backward loop primer
molH2O: molecular grade water
NTC: no-template control
PCM: phase-change material
PCR: polymerase chain reaction
PTC: positive template control
RNA: ribonucleic acid
SOP: standard operating procedure
TTP: time-to-positivity
WHO: World Health Organization

