## Supplemental Materials, Example SOP for "Electricity Free LAMP: A Protocol for Loop-Mediated Isothermal Amplification Using an Exothermic Chemical Reaction and Lyophilized Reagents"

### 1. Materials

- LyoPrime reaction mix (lyophilized, WarmStart-compatible)
- Target-specific LAMP primer set (pre-dispensed or encapsulated)
- Sample nucleic acid (prepared by an appropriate extraction method)
- **Pre-measured reagent microcapsules** (for aqueous rehydration and sample addition)
- Reaction vessels compatible with isothermal amplification
- Insulated thermos or bottle (serving as reaction water bath)
- Aluminum metal, sodium hydroxide solution, sodium chloride (for heat generation)
- Temperature indicator (e.g., thermometer strip or probe)

*Note:* Microcapsules are used in place of micropipettes to reduce user error and eliminate the need for calibrated liquid-handling equipment.

---

### 2. Safety Considerations

Hydrogen gas is generated during the aluminum–base reaction; heating should be conducted in a **well-ventilated area**. Sodium hydroxide is caustic and should be handled using appropriate protective measures. Reaction vessels should remain sealed throughout amplification to prevent contamination.

---

### 3. Preparation of the Chemically Driven Water Bath

A chemically driven heating system is assembled by combining the aluminum metal with aqueous sodium hydroxide and sodium chloride inside an insulated container containing water. The system is **designed to raise water temperature from approximately room temperature**

**(~22 °C) into the LAMP-compatible range (63–67 °C)** and maintain this range for approximately **60 minutes**. For a different starting temperature, refer to Equation 2.

Temperature is monitored to confirm suitability prior to initiating amplification via the temperature indicator, although this is optional.

---

### 4. LAMP Reaction Setup

Suppose a 6 tube setup, each with 25 uL. One tube consisting of the necessary amount of lyophilized is resuspended with 80 uL of input.  3 tubes of the lyophilized LAMP reagents are resuspended with the primer mix, while the other three are resuspended with 25 µL of water.

---

### 5. Isothermal Amplification

Sealed reaction vessels are placed into the chemically heated water bath once the temperature stabilizes within the target range. Amplification proceeds under isothermal conditions for the designated incubation period (~60 minutes).

---

### 6. Result Readout

Following incubation, reactions are removed from the water bath and allowed to cool briefly. Amplification outcomes are assessed using the designated detection modality (e.g., colorimetric change, fluorescence, or turbidity), according to the LyoLAMP system design.

---

### 7. Disposal

Spent heating reagents and reaction vessels are neutralized and disposed of according to institutional chemical and biological waste guidelines.
